# Task-dependent Alteration in Delta Band Corticomuscular Coherence during Standing in Chronic Stroke Survivors

**DOI:** 10.1101/2023.07.17.23292472

**Authors:** Komal K. Kukkar, Nishant Rao, Diana Huynh, Sheel Shah, Jose L. Contreras-Vidal, Pranav J. Parikh

**Affiliations:** Center for Neuromotor and Biomechanics Research, Department of Health and Human Performance, University of Houston, Houston, Texas; Haskins Laboratories, Yale University, New Haven, Connecticut; Laboratory for Noninvasive Brain-Machine Interface Systems, Department of Electrical and Computer Engineering, University of Houston, Houston, Texas

**Author notes:** **Corresponding Author:** Pranav J. Parikh, Ph.D., Department of Health and Human Performance, 3875 Holman Street, suite 104R GAR, University of Houston, Houston, TX 77204, USA, E.

**Keywords:** Stroke, Balance, Corticospinal integrity, EEG, Perturbation, Coupling

## Abstract

Balance control is an important indicator of mobility and independence in activities of daily living. How the changes in functional integrity of corticospinal tract due to stroke affects the maintenance of upright stance remains to be known. We investigated the changes in functional coupling between the cortex and lower limb muscles during a challenging balance task over multiple frequency bands in chronic stroke survivors. Eleven stroke patients and nine healthy controls performed a challenging balance task. They stood on a computerized platform with/without somatosensory input distortion created by sway-referencing the support surface, thereby varying the difficulty levels of the task. We computed corticomuscular coherence between Cz (electroencephalography) and leg muscles and assessed balance performance using Berg Balance scale (BBS), Timed-up and go (TUG) and center of pressure (COP) measures. We found lower delta frequency band coherence in stroke patients when compared with healthy controls under medium difficulty condition for distal but not proximal leg muscles. For both groups, we found similar coherence at other frequency bands. On BBS and TUG, stroke patients showed poor balance. However, similar group differences were not consistently observed across COP measures. The presence of distal versus proximal effect suggests differences in the (re)organization of the corticospinal connections across the two muscles groups for balance control. We argue that the observed group difference in the delta coherence might be due to altered mechanisms for the detection of somatosensory modulation resulting from sway-referencing of the support platform for balance control.

## INTRODUCTION

Poor ability to maintain an upright stance is a significant problem following stroke. It is known to restrict mobility, reduce independence in activities of daily living, and increase the risk of falling during functional tasks (Alenazi et al., 2018; Li et al., 2019; Wong et al., 2016). The control of balance, and its recovery in stroke patients, may depend on changes in the brain regions and the underlying descending pathways (Braun et al., 2007; Fujimoto et al., 2014; Grefkes & Ward, 2014; Mihara et al., 2012; Mima et al., 2001). Standing balance relies on the ability to maintain body’s center of mass (COM) within the base of support (Goel et al., 2019a; Li et al., 2019) by integrating visual, vestibular, and somatosensory inputs (Goel et al., 2019a; Mohapatra et al., 2014). This processing is usually driven by spinal and subcortical regions (Goel et al., 2019a; Sherrington et al., 2017). When the standing balance is challenged, conscious cortical processing is triggered to control lower limb muscles for balance control (Fujimoto et al., 2014a; Hülsdünker et al., 2015; Mihara, Miyai, Hattori, Hatakenaka, Yagura, Kawano, Okibayashi, et al., 2012; Ozdemir et al., 2018a; Slobounov et al., 2006). Following a stroke, the adaptive and maladaptive changes in the cortex and its altered connections with spinal cord/muscles might influence the planning and execution of motor commands for balance compensation. There is a correlation between the integrity of underlying corticospinal pathways and motor function in stroke (Lindenberg et al., 2010; Carlowitz-Ghori et al., 2014; Yoo et al., 2019). How stroke affects the functional integrity of corticospinal tract for the maintenance of upright stance remains to be known.

Corticomuscular coherence is a widely used method to measure corticospinal tract integrity during a functional task (Sinha et al., 2020). In stroke patients, corticomuscular coherence is known to be associated with cortical reorganization (Carlowitz-Ghori et al., 2014) and is linked to functional and behavioral recovery (Grefkes & Ward, 2014; Hara, 2015; Mima et al., 2001; Olafson et al., 2021; Swayne et al., 2008; Wist et al., 2016). Cortico-muscular coherence quantifies the degree of oscillatory coupling (i.e., frequency domain) between the sensorimotor cortex and muscle. In healthy adults, the coherence is primarily seen during challenging standing tasks that require a dynamic control of posture such as while foot stamping and stepping (Masakado Y. & Ushiba J., 2008; Peterson & Ferris, 2019; Stokkermans et al., 2022) but not during quiet standing (Masakado Y. & Ushiba J., 2008). During challenging standing tasks in healthy subjects, corticomuscular coherence is observed in theta and alpha frequency bands for the lower leg muscles such as tibialis anterior and gastrocnemius medialis (Masakado Y. & Ushiba J., 2008; Peterson & Ferris, 2019). Jacobs et al., 2015 found beta band corticomuscular coherence during standing tasks that required changes in stance condition. Consistent with these findings, a recent study (Stokkermans et al., 2023) that used a dynamic standing task requiring production of reactive responses to perturbation at different intensities in forward and backward directions found that the cortex interacts with the leg muscles over a broader frequency spectrum in healthy adults. Mainly, the corticomuscular coherence was observed in all frequency bands there were investigated including theta, alpha, beta, and gamma frequency bands in the lower leg muscles. The coherence in the lowest frequency band, the delta band, has been found to be significant during a similar reactive standing task in older adults (Ozdemir et al., 2016). Following a stroke, the magnitude of corticomuscular coherence has been found to correlate with the recovery in motor function (Grefkes & Ward, 2014; Hara, 2015; Mima et al., 2001; Olafson et al., 2021; Swayne et al., 2008; Wist et al., 2016).

In the chronic stage, several stroke studies have reported changes in coherence between the cortex and the muscles on the affected side during tasks requiring the upper extremity (Braun et al., 2007; Carlowitz-Ghori et al., 2014; Fang et al., 2009; Graziadio et al., 2012; Mima et al., 2001) and knee flexion/extension movements (Ma et al., 2014). The corticomuscular coupling was found either in the beta frequency band (Fang et al., 2009) or over a wider frequency spectrum (Braun et al., 2007), including the gamma frequency band (Ma et al., 2014). However, whether chronic stroke patients demonstrate significant involvement of cortex during quiet standing task and how the coherence modulates when the stance is challenged remain to be known.

In this study, we investigated the changes in the functional coupling between the cortex and the specific lower limb muscles during a challenging balance task over multiple frequency bands in chronic stroke survivors. This data was compared with data from healthy adults in the same age range (healthy controls). We instructed subjects to maintain balance with eyes closed while standing quietly and while we introduced small or large distortions in the somatosensory inputs by altering the gain of a sway-referenced support surface, which influences measures of postural performance, as done in our previous studies (Goel et al., 2019a). The distorted somatosensory inputs acted as a challenge to the upright stance, thus requiring online adjustments of postural movements without which the stability is compromised resulting in a fall. We have earlier shown that varying the gain of the support platform influences spatial and temporal COP-based measures of postural performance, e.g., increase in COP velocity (Goel et al., 2019b, 2021). We simultaneously performed neuroimaging using electroencephalography (EEG) and electromyography (EMG). We hypothesized that stroke patients would demonstrate involvement of the cortex during both quiet standing and challenging balance tasks. We expected that, when compared with healthy controls, stroke patients would demonstrate smaller coherence for the leg muscles over a broader frequency spectrum and that this group difference would be greater during performance of the challenging balance task. The findings from these studies may provide insights into the neural mechanisms underlying poor balance control that may facilitate the assessment of balance behavior during a course of an intervention and design of effective neuromodulation strategies and innovative brain-machine interface-robotics for balance rehabilitation post-stroke.

## METHODS

### Participants

Eleven stroke patients (age: 60.8 ±9.7 years, mean ±SD; range: 37-70; three women) and nine healthy adults (age: 53.6 ±9.4 years, mean ±SD; range: 35-63; four women) provided informed written consent to participate in this study. Demographics are presented in table 1. The inclusion criteria for stroke patients included middle cerebral artery (MCA) stroke for the first time, at least 12 months post stroke, ability to stand for 5 minutes independently without assistance, between 18-90 years of age, have MoCA score ≥ 26 (Bränström et al., 2021; Trzepacz et al., 2015), and absence of other neurological/ musculoskeletal impairments. Healthy adults (controls) were recruited if they had no history/symptoms of neurological/neuromuscular disorders affecting lower limbs with a MoCA score of 26 or greater. Healthy adults were right foot dominant based on self-reporting of the leg with which they preferred to take a step. This study was approved by the Institutional Review Board (00001590) at the University of Houston.

**Table 1:** Clinical information of patients. ^a^ #: with ankle foot orthosis; F, female; M, male; MCA, middle cerebral artery; BBS: Berg Balance Scale; TUG: Timed Up and Go.

| Stroke Patient | Sex | Lesion Site | BBS Score<br>(Total = 56) | TUG Score<br>(sec) |
| --- | --- | --- | --- | --- |
| 1 | M | Right MCA | 44 | 60<sup>#</sup> |
| 2 | M | Right MCA | 44 | 20 |
| 3 | F | Right MCA | 46 | 25 |
| 4 | M | Left MCA | 46 | 24 |
| 5 | F | Left MCA | 53 | 15 |
| 6 | M | Right MCA | 34 | 46.9 |
| 7 | M | Right MCA | 49 | 15.4 |
| 8 | M | Right MCA | 41 | 17.4 |
| 9 | M | Right MCA | 44 | 13.1 |
| 10 | F | Right MCA | 45 | 14.9 |
| 11 | M | Left MCA | 32 | 18.3 |

### Instrumentation

#### Computerized dynamic posturography (CDP)

The dynamic postural stability was assessed using a commercially available CDP Force platform (Neurocom Balance Master, Natus Medical Incorporated, Pleasanton, CA). It is extensively used both in clinical (Cohen & Kimball, 2008) and research settings (Wood et al., 2015) for monitoring sensory and motor performance aspects of the postural control system. The platform is equipped with a motorized dual force plate system (45.72 cm x 45.72 cm), in which ground reaction forces (GRF) from under the subject’s feet are collected by normal and shear force transducers embedded within the force plate (support surface). The system can be used to adjust the orientation of the force plate with respect to the gravitational vertical by rotating it in the sagittal plane about an axis through the subject’s ankle joint in some proportion (a pre-selected gain between −2 to +2) to the postural sway of the subject. A negative sway gain means the movement of the plate will be in the opposite direction of the subject’s COP, and a positive sway gain means the movement of the plate will be in the same direction as the subject’s COP. The transducer data was collected at 100 Hz and processed by pre-installed software on a Windows-based desktop connected to the Neurocom Balance Master (Research module, Neurocom software version 8.0, Natus Medical Incorporated, Pleasanton, CA). The Neurocom system also generated an analog timing signal, which was used to synchronize the electroencephalography (EEG) and the electromyography (EMG) system with the GRF data.

#### Clinical Assessment of balance and mobility

Berg Balance Scale (BBS): The scale consists of a series of 14 predetermined tasks to assess static and dynamic balance. Each task is scored, ranging from 0-4; “0” indicates the lowest functional level, and “4” indicates the highest functional level. The score can take a value between 0 and 56 with higher scores suggestive of better balance (Berg et al., 1992).

Timed Up and Go (TUG) Test: The TUG test measures dynamic balance and functional mobility. The subjects were instructed to rise from an armchair, walk 3 meters from the marker, turn around, walk back to the chair, and sit down. The time to complete this test was measured in seconds. More time taken is associated with poor dynamic balance/mobility (Podsiadlo et al., 1991). Three trials were performed, and a mean score (seconds) was used in subsequent analysis.

#### Laboratory based assessment of reactive balance control

We designed a dynamic balance paradigm, using a continuous balance task, to study cortical involvement in dynamic balance control. The balance control system must continuously process information from multisensory systems to assess postural stability and orientation of the body, and initiate appropriate behavioral responses in case of a disturbance to prevent a fall. During the continuous balance task, participants stood on the balance master with eyes closed while EEG, EMG and GRF data were collected (Fig. 1). The responsiveness of the platform was varied in proportion to the estimated center of mass sway. The sway-referencing of the support surface reduces the contribution of lower limb somatosensory receptors to the control of dynamic balance (Rasman et al., 2018), and thereby manipulated the reliance on somatosensory inputs for the control of ongoing stance and modified the feedback relationship within the balance control loop. Importantly, the effects of such manipulation on dynamic stability are related to the action of the participant. We altered the relationships between postural sway and somatosensory inputs by varying the gain of the support platform in the sagittal plane in different proportions to the estimated instantaneous COM sway angle. We used a range of gains (−1.0, −0.4, 0, 0.4, 0.6, 1.0, 2.0) in the balance task of three different levels of postural difficulty: *low*, *medium* and *high*. The lowest gain value of 0 (a quiet stance with no sway referencing) may not be challenging to subjects and therefore classified as *low* difficulty. Higher gain values of 0.4, 0.6, and 1.0 were classified as *medium* difficulty as subjects were expected to exhibit greater postural instability. However, the gain of 2.0, or negative gains of −0.4 and −1.0 were classified as that of *high* difficulty due to expectations that subjects would have a further increase in postural instability, as done in our previous work (Goel et al., 2019a; Ozdemir et al., 2016, 2018b).

**Fig. 1.**
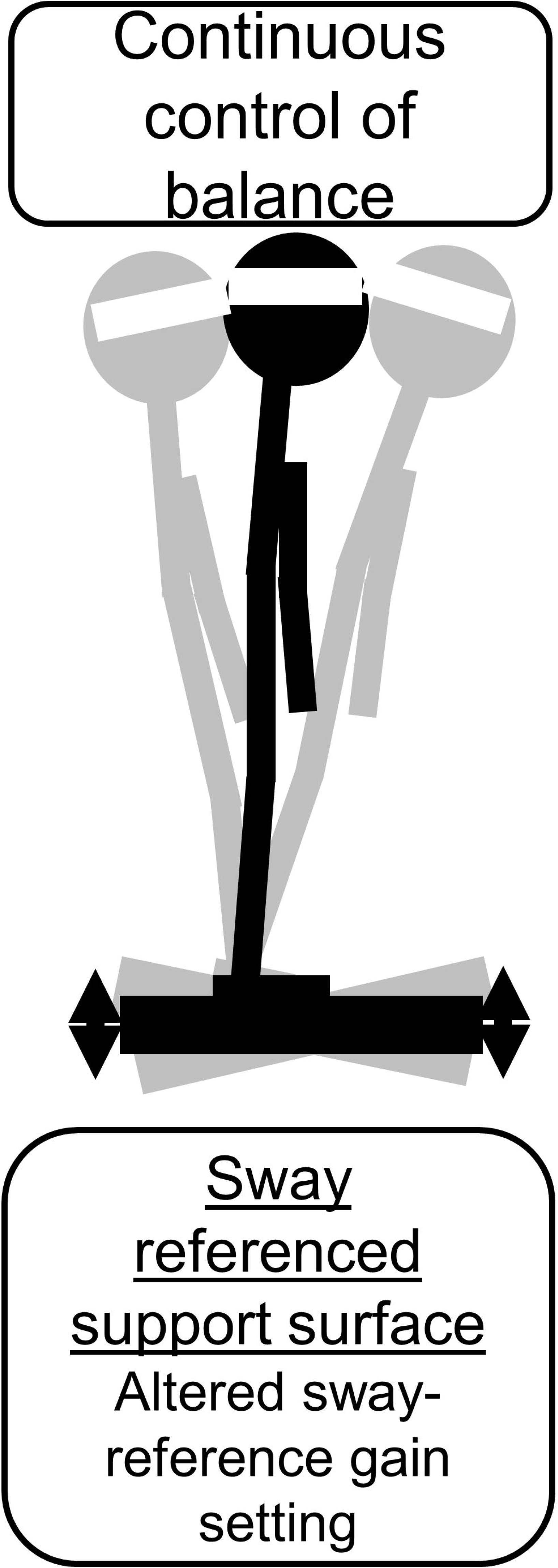
Experimental setup and the continuous balance task. **A.** Experimental setup for the posture task. **B.** Continuous balance task. Schematic description of experimental conditions during the continuous balance task.

The continuous balance task lasted for 180 s, with three 20 s testing conditions at a gain of 0 and one 20 s condition for each of the other four gain values presented in the following order for each subject: 0, 0.6, −0.4, 0, 1.0, −1.0. During the task, subjects were instructed to maintain their balance. The order of these conditions was chosen so that we first had a *low* difficulty condition with a stable support surface (gain value of 0), and then the difficulty was gradually increased to *medium* level (gain values of 0.6 or 1.0), and then increased further to *high* level (gain values of - 0.4 or −1.0). The general pattern of *low*, *medium*, and *high* difficulty was repeated three times. These varying postural conditions allowed us to gradually increase the difficulty of the postural control task while simultaneously monitoring the EEG, EMG, and GRF responses. Thus, this task was developed with the specific intent of challenging the postural control system in such a way that both its behavioral (GRF measures) and neuromuscular (EEG and EMG measures) underpinnings could be observed during the stable (low difficulty), cautious (medium difficulty), and threatened (high difficulty) stages of balance control. Subjects were not informed when the gain changed. To ensure safety, subjects always wore a safety harness. In addition, they also wore a physical therapy belt with a spotter standing nearby to prevent any falls. Subjects were allowed to practice at gain values of 0, 0.4, 2.0 prior to exposure to the start of the data collection. Data from these gain values was not used for analysis.

#### Electroencephalography (EEG)

Whole-scalp electroencephalography (EEG) was recorded using 64 active channel EEG electrodes (Brain Products GmbH, Germany; 1000 Hz). Four of these electrodes were used to record electrooculography (EOG) signals. EEG was recorded at rest and during the balance task. We constrained movement of the EEG cables by placing an elastic mesh on the top of the EEG cap, which aided in minimizing motion artifacts during the balance task on the Neurocom (Goel et al., 2019a; Luu et al., 2017). We utilized a modified international 10-20 EEG system where we moved the GND and REF from the default locations (AFz and FCz) to the left and right earlobes, respectively (Luu et al., 2016). We made these modifications because the default channel locations for GND and REF are very close to the fronto-central cortex, a region of interest for this study. The empty spots were filled by moving T7 and T8 electrodes to AFz and FCz locations, respectively.

#### Electromyography (EMG)

We recorded activity of thigh and leg muscles bilaterally using differential surface electrodes from tibialis anterior (TA), medial gastrocnemius (GM), soleus (SOL), rectus femoris (RF) and biceps femoris (BF) (1111.11 Hz, gain 1000; Delsys Trigno EMG System, Boston, MA). These muscles were selected because of their involvement in the control of vertical posture while dealing with symmetrical perturbations induced in the sagittal plane (Mohapatra et al., 2014; Santos et al., 2010). The placement of electrodes for recording EMG activity was based on recommendations reported in the literature (Mohapatra et al., 2014).

### Experimental Protocol

Each patient and healthy control participated in a single session. First, we obtained their clinical measures using BBS and TUG test. Following this step, we prepared subjects for EEG and EMG measurements. Prior to electrode placement, the skin was prepared by cleaning with isopropyl alcohol pads and by shaving excessive hair, if necessary. Each subject then performed the challenging balance task with varying sensory conditions while standing on the force platform. We obtained baseline measurements for 60 sec during quiet standing with eyes closed before and after the balance task. The dynamic balance task consisted of maintaining an upright stance with eyes closed during a nine sequential 20 s duration posture test conditions.

### Data Analyses

#### Behavioral data

BBS (out of 56) and TUG (average of three trials in seconds) were collected by the physical therapist before the balance task for further analysis. The ground reaction force data collected from the force plate were combined to create a center of pressure (COP) time series in medio-lateral (ML) and anterior-posterior (AP) directions for the continuous balance task (Neurocom, 2009). Next, we estimated the COM by low pass filtering the COP data (second order Butterworth; f_c_ = 0.86 Hz) (Goel et al., 2019a, 2021) for the AP direction for the continuous balance task as the sway of interest was in the AP direction due to the mechanical configuration for rotation of the support surface. Several studies have confirmed the reliability of this method to estimate COM from COP (Breniere, 1996; Caron et al., 1997; Lafond et al., 2004).

For the continuous balance task, we computed root mean square (RMS) COP (Prieto et al., 1996) and path length (PL) (Donath et al., 2012) from COP (Forth et al., 2011; Ozdemir et al., 2013, 2018b). The RMS and PL of COP represent spatial measures of postural performance. We also calculated COP velocity (RMSCOPv) (Geurts et al., 1993), which is also a reliable measure in the quantification of postural control.

The number of falls induced by the platform translations was also recorded using qualitative observations during testing. A fall was defined as a participant who lost their balance and required either self-induced stepping and/or the overhead harness to prevent them from falling to the floor (e.g., participants applied force through the rope and harness system as evident from the rope and harness going taut). The spotter was instructed to assist the participant only after they had fallen (i.e., to prevent swinging caused by the rope).

#### EEG pre-processing

The EEG pre-processing used in this study was similar to that described in (Goel et al., 2018a). Briefly, the raw EEG signals sampled at 1000 Hz were first downsampled to 250 Hz. EOG channels (4 channels) were utilized for adaptive filtering of the EEG signals (60 Channels) through the H infinity algorithm to remove eye artifacts, correct baseline drifts, and remove other shared sources of noise (Kilicarslan et al., 2016). A standardized early-stage EEG processing pipeline (PREP) with default parameters was used to remove artifactual EEG channels and apply a robust common referencing method to increase the signal to noise ratio (Bigdely-Shamlo et al., 2015). PREP is reported to be more robust in detecting noisy channels compared to the previously known noisy channel detection method: *pop_rejchan* from EEGLAB (Bigdely-Shamlo et al., 2015; Delorme & Makeig, 2004). The PREP pipeline also replaces the artifactual channels with surrogate data that is interpolated from neighboring sensors. This minimizes a bias when performing common average referencing. The signals were then band pass filtered at 0.1-100 Hz using a 4^th^ order Butterworth filter to remove slow drifting noise. Artifact Subspace Reconstruction (ASR) was applied next to detect and denoise any artifactual sections in the EEG data (Mullen et al., 2013). The ASR uses clean EEG data to calibrate the noise covariance and uses a standard deviation threshold of 15 with a sliding window (500 ms) based principal component analysis (PCA) to detect noisy sections in the EEG data (Artoni et al., 2017; Chang et al., 2020; Mullen et al., 2013).

Next, to compensate for the rank deficiency in the data, surrogate channels during the PREP pipeline were removed before running the independent component analyses (ICA) since the ICA assumes the number of independent components to be the same as the number of channels provided as the data. Adaptive mixture ICA (AMICA) was used to compute the maximally independent components (ICs) from the data (Palmer et al., 2011). The AMICA is reported to be the best ICA method available to date, and it provides more dipolar and minimized mutual information ICs (Delorme et al., 2012; Leutheuser et al., 2013). Using the constructed boundary elemental model (BEM) explained in the next paragraph and the digitized channel locations, dipole fitting method: *DIPFIT* in EEGLAB (Delorme et al., 2011; Delorme & Makeig, 2004) was used to calculate the equivalent current dipole sources that explain at least 85% (Sipp et al., 2013) of topographic variance obtained from ICA results. The digitized channel locations were first warped onto the constructed head model before calculating the dipole locations. We removed dipoles located outside the individual BEM model as well as those with artifactual components such as muscle-related power spectral density characteristics and motion artifact related high-frequency noise.

The boundary element model (BEM) was created after computing the surface meshes and constructing the head model using a standard MRI template. The constructed BEM was then saved and used as one of the arguments in the *DIPFIT* process. Each IC scalp projection, its equivalent dipole’s location, and its power spectra were then visually inspected and ICs that related to non-brain artifacts (e.g., sensor movement, muscle artifact, stimulation artifact) were removed.

### Wavelet coherence analysis

Wavelet coherence (WC) was used to investigate the relation in time-frequency space between the EEG and EMG signals. It is a method of measuring the cross-correlation between two signals with respect to time and frequency. We computed WC between Cz (EEG) electrode and EMG activity measured from TA, SOL, GM, BF, and VL using signals from the affected side in stroke patients and from the dominant side in healthy adults. In absence of age- and gender-matching between the two groups, it was not appropriate to study the corresponding side in healthy controls. We used the Morlet wavelet as the mother wavelet, with default parameters set as one octave for 10 scales as it provides good time and frequency localization, and a complex shape that allows for capturing both amplitude and phase information (Grinsted et al., 2004). The localized coherence significance was evaluated using Monte Carlo analysis with 1000 iterations as this provide a way to estimate statistical significance and assess the reliability of the wavelet results. EMG signals were detrended and downsampled to 250 Hz. The final two seconds of the EMG signal was corrupt, and this was found across several trial conditions and thus this was eliminated from further analysis. WC was computed over the first 18 sec of trial (4500 samples) with a frequency resolution of 0.0526 Hz. To understand the dynamic nature of coherence between EEG and EMG, WC was averaged over 2 sec non-overlapping trial segment (9 bins). Values outside the cone of influence (COI) were considered highly non-reliable and thus excluded from the analysis(Grinsted et al., 2004). This also led to the exclusion of the first and last bins. Coherence was considered significant (p < 0.05) if it was greater than Z, which is given using the following formula:

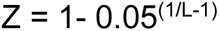

where L is the total number of non-overlapping segments, equal to the available number of trials multiplied by six (Witham et al., 2011). The average coherence values were then converted to Fisher z-statistics (normalized) using arc hyperbolic tangent transformation (M. R. Baker & Baker, 2003a; Mima et al., 1999) prior to performing any statistical analysis. We averaged coherence spectra across trials and groups to calculate peak coherence and associated peak frequency for both the groups.

The Cross wavelet and Wavelet Coherence Package (Grinsted et al., 2004) also provides time-phase direction, shown by the orientation of arrows in Fig. 3. Arrows pointing straight right demonstrates perfect in phase while arrows pointing straight left represents perfect out of phase relationship. In addition, arrows pointing downward represents EEG leading EMG by 90 degrees whereas straight up arrow shows EMG leading the EEG by 90 degrees.

### Statistical Analyses

For behavioral, EEG, and EMG data, we performed normality (Shapiro-Wilk) and equal variance Levene’s test to check for violation of assumptions for parametric statistical approach. For BBS and TUG scores, these assumptions were found to be violated (both P values > 0.05) and thus we used a non-parametric approach, Mann-Whitney U test, to compare the stroke group with the healthy group. The assumptions were validated for the COP, EEG, and EMG data.

For RMS COP, RMS COP_v_ and COP PL, we used restricted maximum likelihood linear models with between-subject factor as Group (Stroke, Healthy) and within-subject factors such as Task Condition (Low, Medium, High). Statistical analysis was conducted on coherence values obtained over the 14-sec trial window for each task condition.

For normalized peak coherence and peak frequency measures, we used separated restricted maximum likelihood linear models for each of the five muscles with between-subject factor as Group (Stroke, Healthy) and within-subject factors such as Task Condition (Low, Medium, High). Next, we obtained average coherence values for each frequency bands: delta (0.1 – 4 Hz), theta (4 – 5 Hz), alpha (8 – 12 Hz), beta (15 – 30 Hz), and low gamma (30 – 50 Hz) (Goel et al., 2019a).

For normalized EEG-EMG coherence measure for frequency bands and bins, we used restricted maximum likelihood linear models with between-subject factor as Group (Stroke, Healthy) and within-subject factors such as Task Condition (Low, Medium, High), Frequency Bands (Delta, Theta, Alpha, Beta, Gamma), and Time Bins (Bin1, Bin2, Bin3, Bin4, Bin5, Bin6, Bin7) for each of the five muscles. The 1^st^ and the last bins were deleted as they were outside the COI. For each statistical model, post-hoc analysis was performed for significant results using Bonferroni correction for multiple comparisons. Further, effect sizes (in terms of Cohen’s d) are provided for paired comparisons where statistical tests showed a significant difference. Cohen’s d value less than 0.20 indicates no noticeable effect, between 0.20 and 0.49 indicate small effect, between 0.50 and 0.79 indicate medium effect, between 0.80 and 1.29 indicate large effect, and above 1.30 indicate very large effect (Lakens, 2013). The α significance level was set at 0.05. Our initial analysis showed no difference in the COP and coherence values between two low, two medium, and two high conditions (separate paired t-test; all P values > 0.05). Therefore, we considered data averaged across gain values of similar level of difficulty for further analysis (e.g., data averaged across two low, two medium, and two high conditions). All average statistics are displayed in the form of mean ± standard error of measurement.

## RESULTS

All subjects in the stroke and healthy groups were able to complete the clinical assessment and the continuous balance task. There was no difference between the two groups for age (t_18_ = 1.622; p =0.122). Subjects in both groups found the high trial condition of continuous balance task to be highly challenging, i.e., subjects lost their balance and required the harness to prevent them from falling to the floor (e.g., subjects applied force through the rope and harness system as evident from the rope and harness going taut). These trials were noted during data acquisition and all data for the entire trial were excluded from data analysis. There was no significant difference between the two groups in the total number of trials eliminated (11 and 6 of 66 trials were eliminated in stroke and healthy control groups, respectively) (χ^2^ (df=1) = 0.321; *p* > .05).

### Balance and mobility performance

#### Performance on clinical tests of balance and mobility

As expected, stroke patients showed poor scores on clinical tests of balance and mobility when compared to healthy controls. Stroke patients (55.1 ± 0.93) when compared to healthy controls (43.5 ± 6.04) showed lower scores on the Berg Balance Scale (Fig. 2; Mann Whitney U-test; *U* = 0.5, *Z* = – 3.765; *p* < .001). Consistent with this finding, stroke patients (24.5 ± 15.05 sec) when compared to healthy controls (8.6 ± 1.1 sec) took longer to complete the timed-up and go task (Fig. 2; Mann Whitney U -test; *U* = 0.0, *Z* = – 3.762; *p* < .001).

**Fig. 2.**
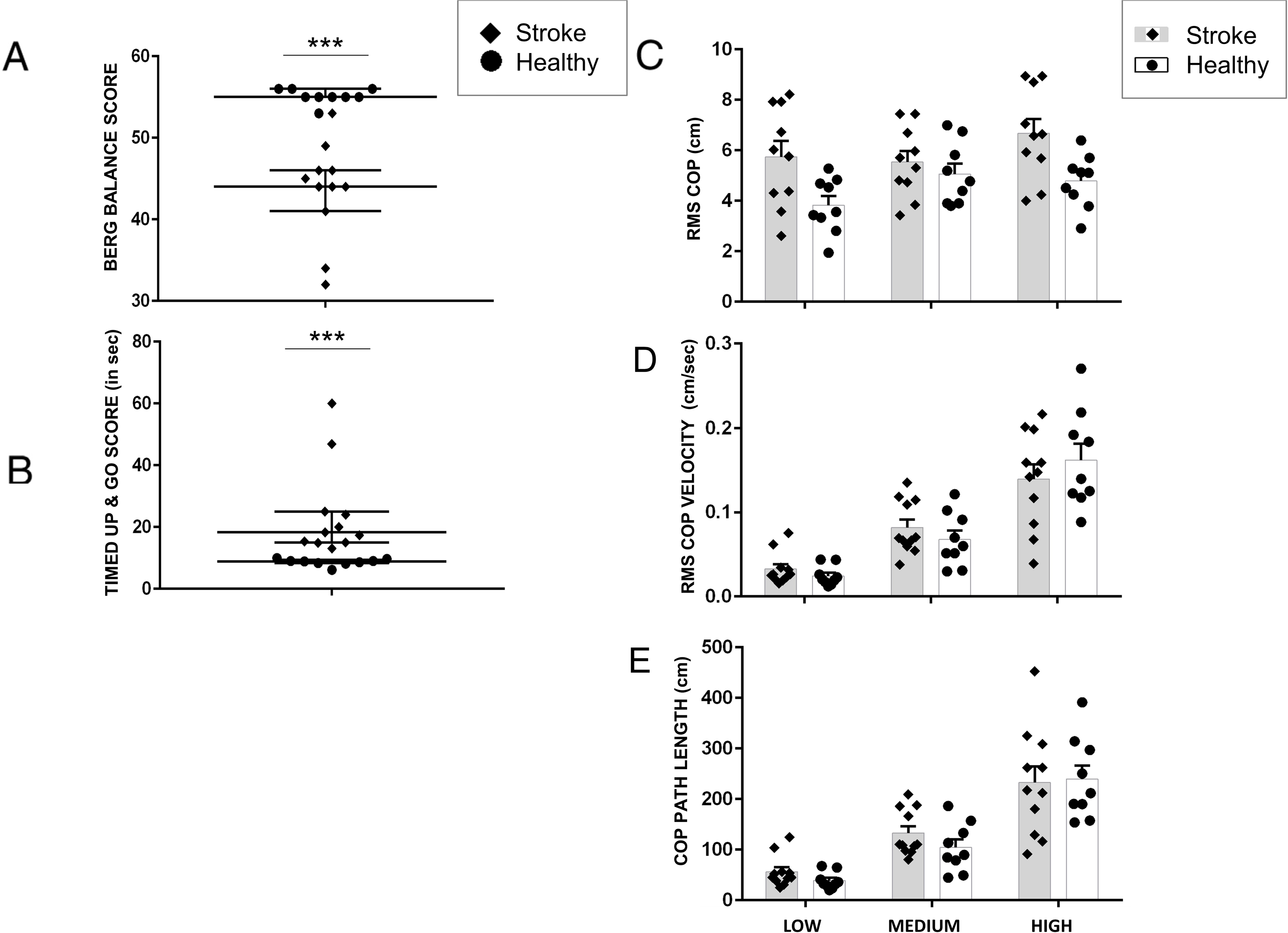
Behavioral variables. **A.** Berg Balance Score (BBS), **B.** Timed Up and Go (TUG), **C.** RMS COP, **D**. RMS COPv, and **E.** COP Path length (PL) during the continuous balance task. Data are means (± SEM) of all subjects. *** P < 0.001., significant between-group differences.

#### Performance on the continuous balance task

Stroke patients and healthy adults performed a continuous balance task under varying sensory conditions. We found that the continuous balance task challenged the postural control system of stroke patients more than healthy controls. Mainly, stroke patients showed larger RMS COP when compared with healthy adults (main effect of Group: F_1,51_ =13.22, p < 0.01) for all conditions (no Group × Condition interaction: F_2,51_ =1.459, p = 0.2419; no main effect of Condition: F_2,51_ =1.933, p =0.1552; Fig. 2).

Although increasing the gain of support surface increased COP PL (main effect of Condition: F_2,54_ = 45.37, p < 0.0001), we found that this increase in PL was similar for the stroke group and the healthy group across difficulty conditions (Fig. 2; No Group × Condition interaction: F_2,54_ = 0.4193, p = 0.6596 and no main effect of Group: F_1,54_ = 0.6293, p = 0.4311). Similarly, we found an increase in RMS COP_v_ with an increasing difficulty of the balance task for both groups (Fig. 2; main effect of Condition: F_2,54_ = 49.44, p < 0.0001). However, this increase in RMS COP_v_ was similar for the stroke group and the healthy group (no significant Group x Condition interaction: F_2,54_ = 1.270, p = 0.2891; no main effect of Group: F_1,54_ = 0.000, p = 0.997). Post-hoc tests for COP-PL and RMS COP_v_ did not reveal any significant differences across conditions.

### EEG – EMG Coherence

Fig. 3 shows a representative example of EEG-EMG coherence spectra for one stroke patient and one healthy adult. The spectral distribution of the EEG-EMG coherence showed the highest significant coherence between EEG over the frontocentral Cz electrode location and EMG of the tibialis anterior or TA muscle of the affected side for stroke patients and the right dominant side for healthy adults. The highest coherence in the stroke group was mostly in medium and high conditions, and for healthy controls it was in medium conditions.

**Fig. 3.**
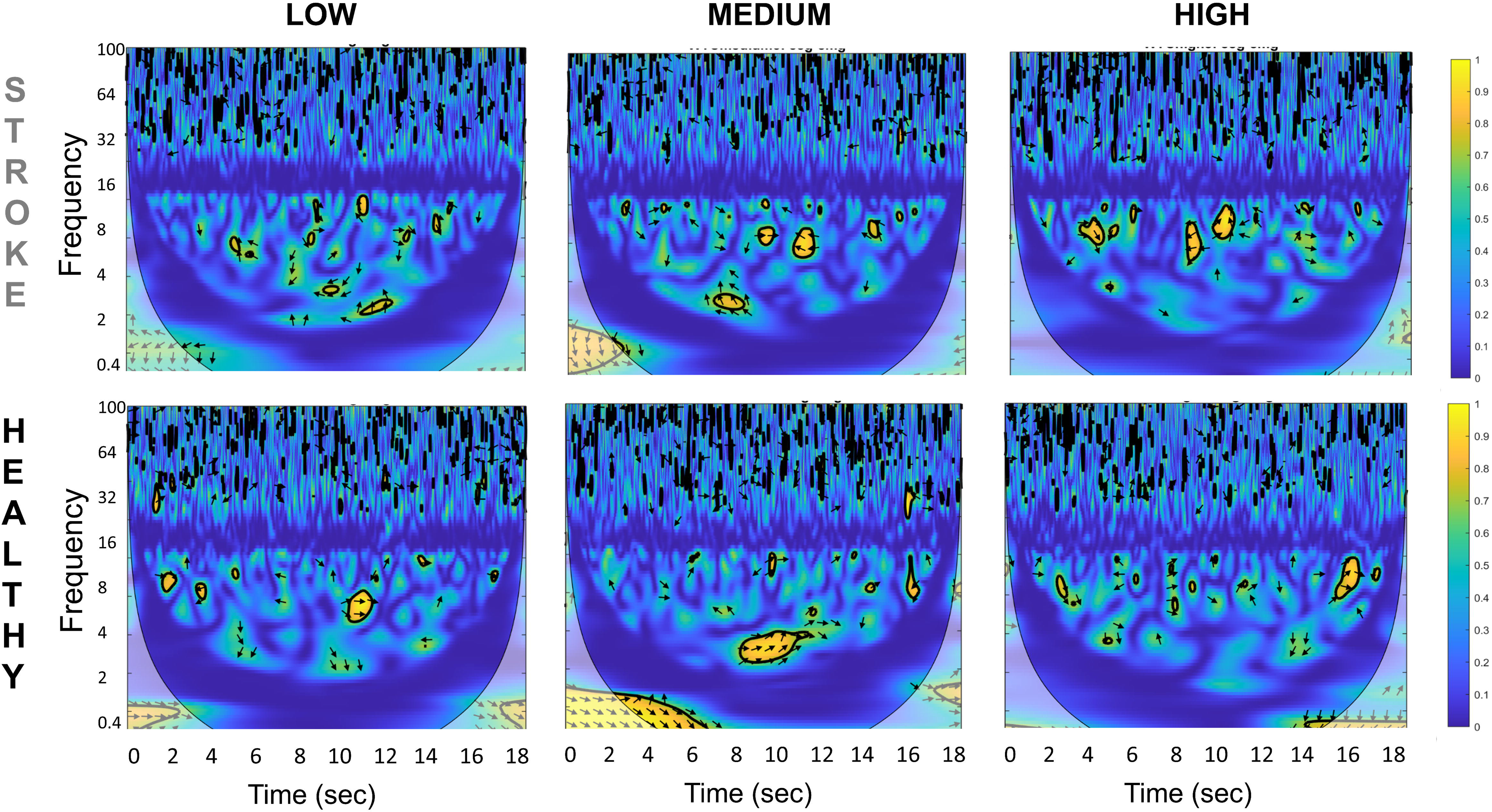
Representative time-frequency cortico-muscular coherence (CMC) plot for one stroke patient (*top row*) and one healthy control (*bottom row*) for all three task conditions (low, medium and high) between Cz EEG electrode and Tibialis Anterior EMG of the affected side.

#### Peak coherence magnitude and Frequency

We first computed the peak coherence magnitude as the highest absolute EEG-EMG coherence value during a trial for both the groups across all three conditions (low, medium and high) and for five muscles (GM, RF, BF, TA, SOL). For GM, the stroke group showed significantly different magnitude of peak coherence when compared with the healthy group and this difference varied across task conditions (F_2,36_ = 5.325, p=0.009; Fig. 4). Post-hoc pair-wise comparisons found that the stroke group showed significantly lower magnitude of peak coherence when compared with healthy group (t_18_ = 3.1591; p = 0.0054) for the medium continuous balance task condition. However, no group difference was found for the low (t_18_ = 1.967; p =0.065) and high (t_18_ = 0.722; p =0.479) conditions.

**Fig. 4.**
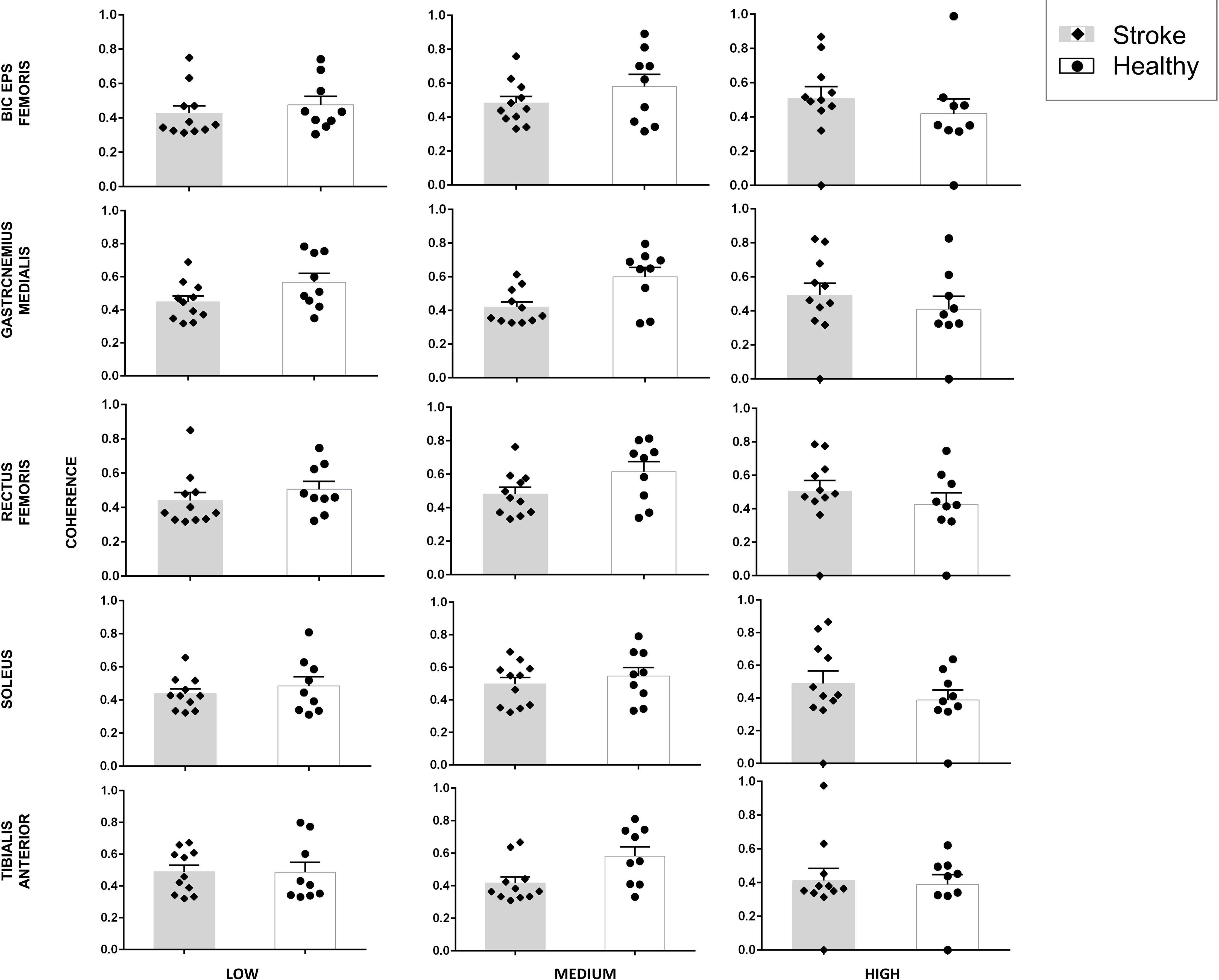
Peak cortico-muscular coherence (CMC) magnitude data for stroke patients (*grey bars*) and healthy control (*white bars*) for all three task conditions (low, medium and high) and for all five muscles of the affected side. Data are means (± SEM) of all subjects.

Similarly, for RF, the stroke group showed significantly different magnitude of peak coherence when compared with the healthy group and this difference varied across task conditions (F_2,36_ = 4.731, p=0.015; Fig. 4). However, post-hoc analysis did not reveal significant group differences for all task conditions (all p values > 0.0167). For BF, no significant results were obtained (no main effect of Group: F_1,18.434_ = 0.704, p=0.412; no main effect of Condition: F_2,34.961_ = 1.833, p=0.175; and no Group × Condition interaction: F_2,34.961_ = 0.216, p=0.807; Fig. 4).

For SOL, no significant results were obtained (no main effect of Group: F_1,18.002_ = 0.008, p = 0.931; no main effect of Condition: F_2,36_ = 0.617, p = 0.545; and no significant Group × Condition interaction: F_2,36_ = 2.588, p=0.089; Fig. 4). For TA, no significant results were obtained (no main effect of Group: F_1,18_ = 0.031, p=0.861; no main effect of Condition: F_2,36_ = 0.044, p = 0.957; and no significant Group × Condition interaction: F_2,36_ = 1.895, p = 0.165; Fig. 4).

Across all subjects in the stroke and healthy groups, the magnitude of EEG-EMG coherence peaked in delta or theta frequency range (Fig. 5) for all muscles. For some muscles, a 2^nd^ peak was also evident especially for high conditions in beta and gamma bands. We did not find any significant differences across groups and conditions for all the muscles (all p values > 0.05) (BF: group- F_1,18.273_ = 1.554, p=0.228, conditions- F_2,34.858_ = 0.227, p=0.798, group*conditions- F_2,34.858_ = 1.393, p=0.262; GM: group - F_1,18_ = 0.019, p=0.892, conditions- F_2,36_ = 0.256, p=0.776, group*conditions- F_2,36_ = 1.492, p=0.238; SOL: group - F_1,18.008_ = 0.032 p=0.860, conditions- F_2,36.039_ = 0.226, p=0.799, group*conditions- F_2,36.039_= 0.514, p=0.602; TA: group - F_1,18_ = 0.093, p=0.764, conditions- F_2,21.108_ = 1.734, p=0.201, group*conditions- F_2,21.108_ = 0.586, p=0.565), except for a significant interaction effect for rectus femoris muscle (RF: group - F_1,18.012_ = 0.982, p=0.335, conditions- F_2,36.016_ = 0.781, p=0465, group*conditions- F_2,36.016_ = 3.976, p=0.028), but post-hoc analysis did not show any significant comparisons for RF muscle.

**Fig. 5.**
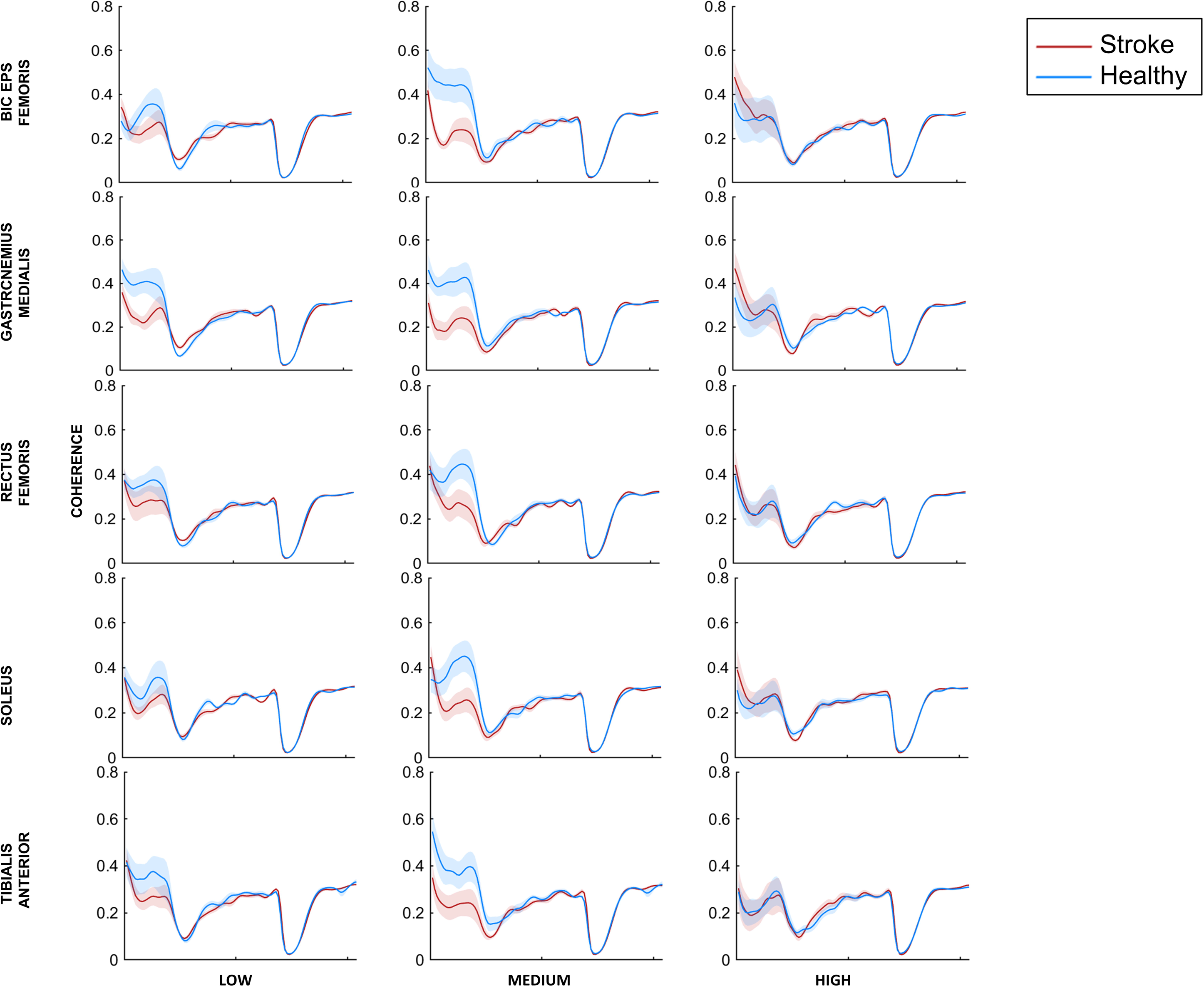
Cortico-muscular coherence (CMC) frequency data for stroke patients (*red line*) and healthy controls (*blue line*) for all three task conditions (low, medium and high) and for all five muscles. Data are means (± SEM) of all subjects.

#### Coherence for frequency bands and trial bins

For TA muscle, we found that increasing the gain of the support surface altered normalized EEG-EMG coherence (main effect of Condition: F_2,5205.6_ =7.739; p <0.001). Mainly, the coherence was greater for the medium difficulty condition when compared to high (medium vs. high mean difference=0.008; p<0.001) and for the low difficulty condition as compared to high (low vs. high mean difference=0.007; p=0.005) difficulty conditions (Fig. 6). Similarly, within a trial, coherence also modulated across frequency bands (main effect of Bands: F_4,5204.5_=4241.7; p<0.001) and bins (main effect of bins: F_6,5203.5_=12.985, p<0.001). Importantly, across the stroke and healthy control group, there was modulation in normalized EEG-EMG coherence across difficulty conditions (significant Group × Condition interaction: F_2,5205.9_ =6.493, p = 0.002) and frequency bands (significant Group × Bands interaction: F_4,5203.9_ =25.785, p<0.001), but not across bins (no significant Group × Bins interaction: F_6,5203.9_ =1.228, p = 0.289; no main effect of Group: F_1,50137640.2_ = 0.055, p =0.815). Pair-wise comparisons showed that stroke patients showed significantly smaller coherence than healthy controls during the medium difficulty condition in the delta frequency band (t_18_ =3.08, p=0.0053; Fig. 6). All other comparisons were non=significant (p>adjusted α for multiple comparisons).

**Fig. 6.**
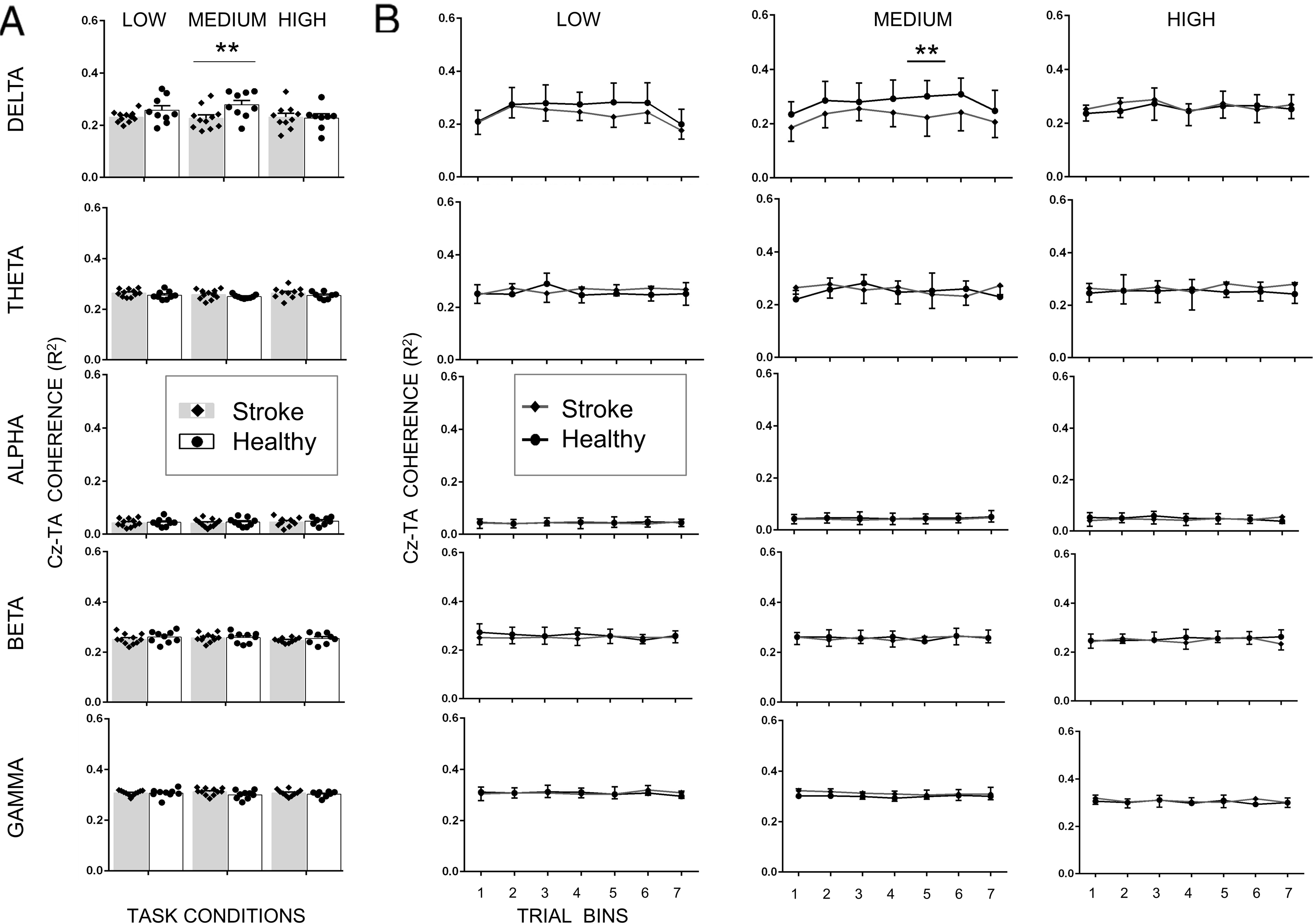
Cortico-muscular coherence (CMC) magnitude between Cz (EEG) and tibialis anterior (EMG) for all five frequency bands and all three task conditions for stroke patients and healthy controls. **A.** CMC averaged across the entire 20 sec trial. **B.** CMC for seven trial bins. Data are means (± SEM) of all subjects. **P < 0.01, significant between-group differences.

For SOL muscle, we found that increasing the gain of the support surface altered normalized EEG-EMG coherence (main effect of Condition: F_2,5136.1_ = 3.781; p = 0.023). Mainly, the coherence was greater for the medium difficulty condition when compared to high (medium vs. high mean difference = 0.006; p = 0.032) difficulty condition. The coherence was also greater for medium difficulty condition than low but did not reach statistical significance (medium vs. low mean difference=0.005; p=0.103) (Fig. 7). The coherence also modulated across frequency bands (main effect of Bands: F_4,5133.7_=4271.9; p<0.001) and bins (main effect of Bins: F_6,5133.7_=11.227, p<0.001). Importantly, across the stroke and healthy control groups, there was a modulation in normalized EEG-EMG coherence across difficulty conditions (significant Group × Condition interaction: F_2,5136.1_ = 12.02, p<0.001) and frequency bands (significant Group × Bands interaction: F_4,5133.7_ =12.306, p<0.001), but not across bins (no significant Group × Bins interaction: F_6,5133.7_ =1.847, p = 0.086; no main effect of Group: F_,1,51010193.1_ = 0.237, p = 0.626). Pair-wise comparisons showed that stroke patients showed significantly smaller coherence than healthy controls during the medium difficulty condition in the delta frequency band (t_18_ =3.854, p=0.0012; Fig. 7). All other comparisons were non=significant (p>adjusted α for multiple comparisons).

**Fig. 7.**
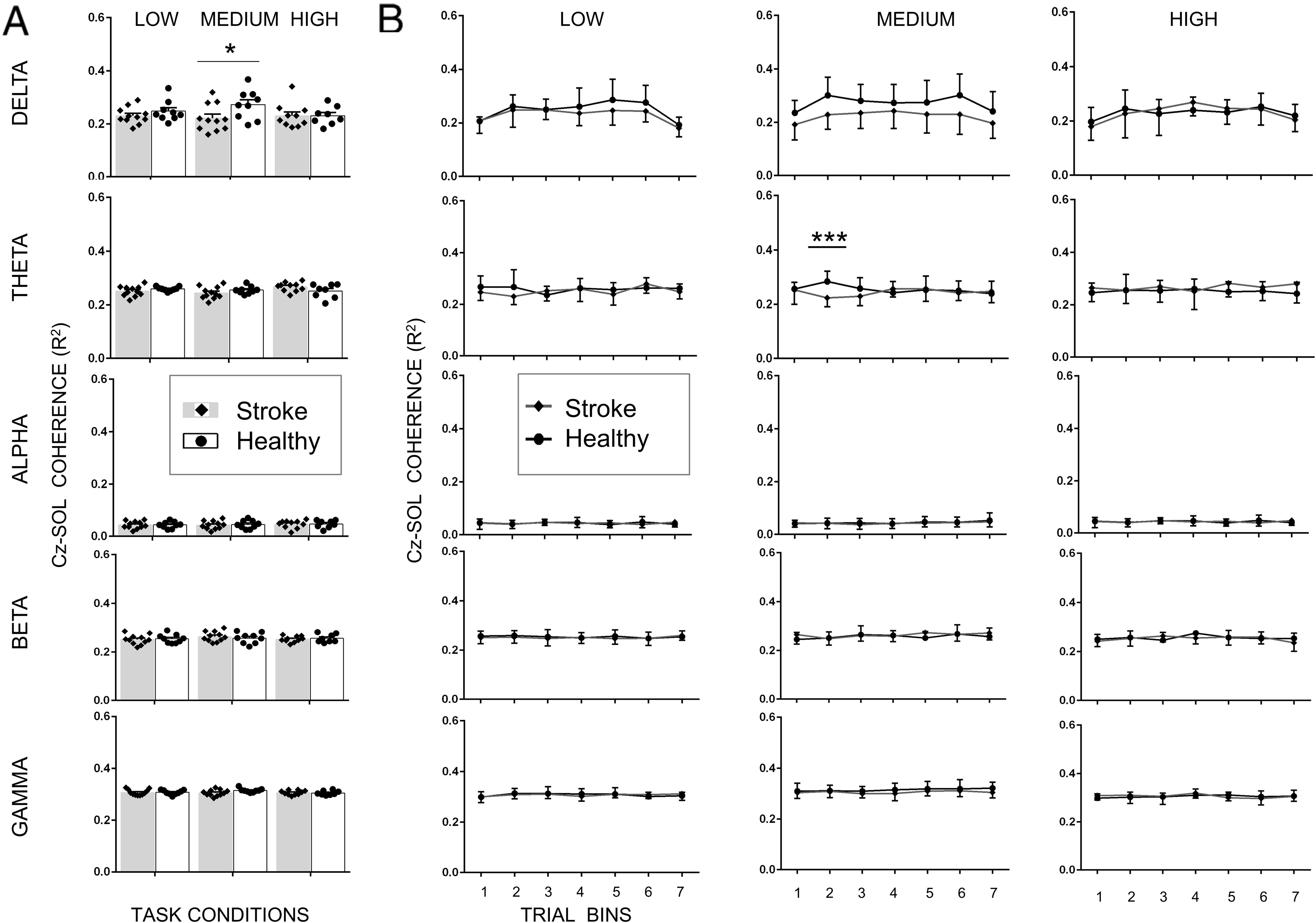
Cortico-muscular coherence (CMC) magnitude between Cz (EEG) and soleus (EMG) for all five frequency bands and all three task conditions for stroke patients and healthy controls. **A.** CMC averaged across the entire 20 sec trial. **B.** CMC for seven trial bins. Data are means (± SEM) of all subjects. **P < 0.01, significant between-group differences.

For GM muscle, we found that within a trial, normalized EEG-EMG coherence modulated across frequency bands (main effect of Bands: F_4,5093.6_=3951.9; p<0.001) and bins (main effect of Bins: F_6,5093.6_=15.287, p<0.001). But, increasing the gain of the support surface did not alter normalized EEG-EMG coherence (no main effect of Condition: F_2,5096.1_ =1.247; p = 0.287) (Fig. 8). But, across stroke and healthy controls, there was a modulation in normalized EEG-EMG coherence across difficulty conditions (significant Group × Condition interaction: F_2,5096.1_ = 3.725 p = 0.024), frequency bands (significant Group × Bands interaction: F_4,5093.7_ =14.18, p < 0.001) and bins (significant Group × Bins interaction: F_6,5093.7_ = 3.429, p = 0.002). Pair-wise comparisons showed that stroke patients showed significantly smaller coherence than healthy controls during the medium difficulty condition in the delta frequency band (t_18_ =3.854, p=0.0012; Fig. 8). All other comparisons were non-significant (p>adjusted α for multiple comparisons).

**Fig. 8.**
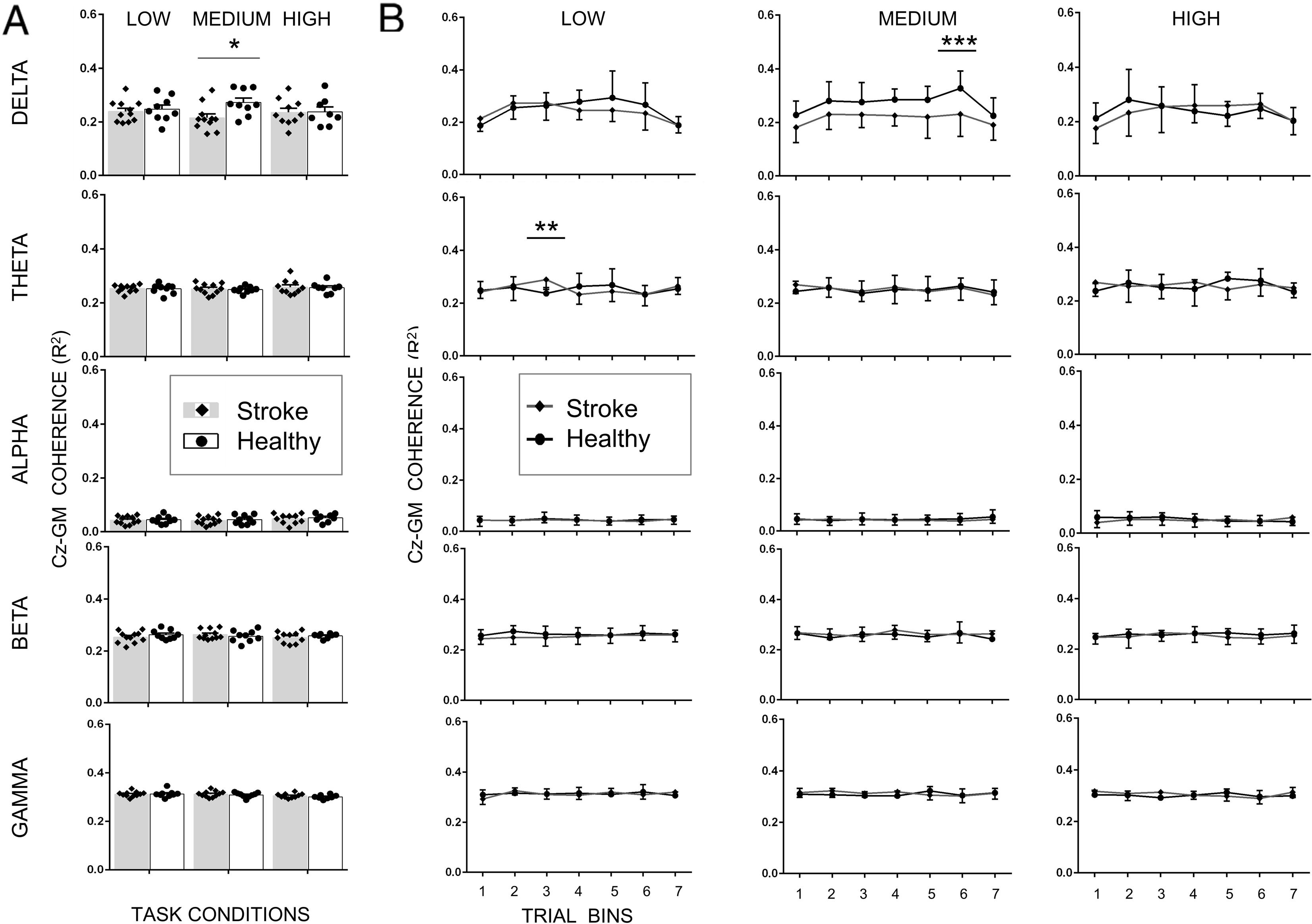
Cortico-muscular coherence (CMC) magnitude between Cz (EEG) and gastrocnemius medialis (EMG) for all five frequency bands and all three task conditions for stroke patients and healthy controls. **A.** CMC averaged across the entire 20 sec trial. **B.** CMC for seven trial bins. Data are means (± SEM) of all subjects. **P < 0.01, significant between-group differences.

For the BF muscle, we found that increasing the gain of the support surface altered normalized EEG-EMG coherence (main effect of Condition: F_2,1811.2_ = 9.596; p < 0.001). Mainly, the coherence was greater for the medium difficulty condition when compared to low (medium vs. low mean difference=0.013; p < 0.001) and high (medium vs. high mean difference = 0.013; p = 0.002) difficulty conditions (Fig. 9). The coherence also modulated across frequency bands (main effect of Bands: F_4,1802.4_ = 2047.64; p<0.001) and bins (main effect of Bins: F_6,1802.4_ = 4.279, p<0.001). Importantly, across the stroke and healthy control group, there was modulation in normalized EEG-EMG coherence across difficulty conditions (significant Group × Condition interaction: F_2,1811.2_ =,3.797 p = 0.023) and frequency bands (significant Group × Bands interaction: F_4,1802.4_ =7.778, p<0.001), but not across bins (no significant Group × Bins interaction: F_6,1802.4_ =0.215, p = 0.972; no main effect of Group: F_,1,18.4_ = 0.849, p =0.369; Fig. 9). All pairwise posthoc comparisons were non-significant (p>adjusted α for multiple comparisons).

**Fig. 9.**
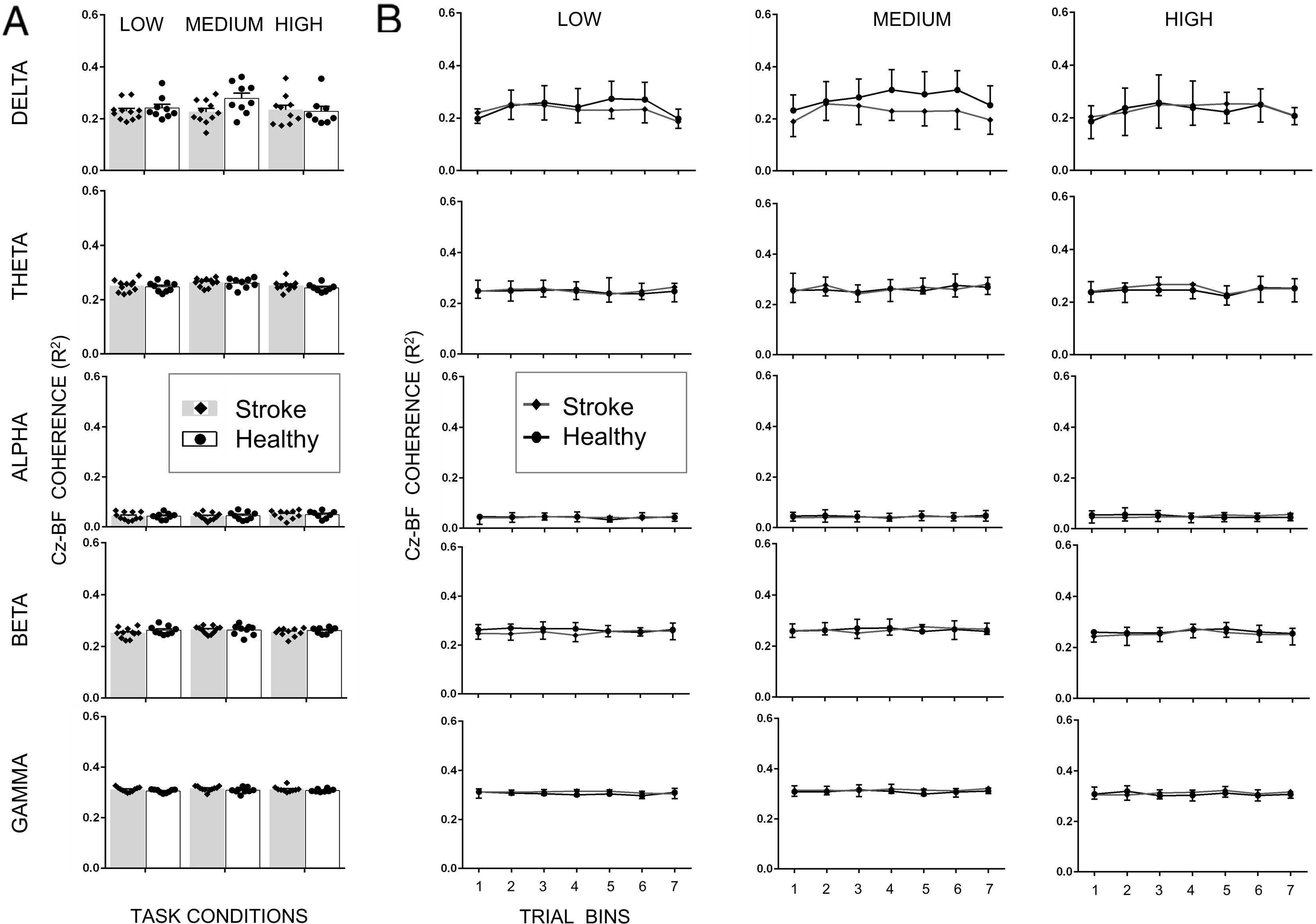
Cortico-muscular coherence (CMC) magnitude between Cz (EEG) and biceps femoris (EMG) for all five frequency bands and all three task conditions for stroke patients and healthy controls. **A.** CMC averaged across the entire 20 sec trial. **B.** CMC for seven trial bins. Data are means (± SEM) of all subjects. **P < 0.01, significant between-group differences.

Similarly, for RF muscle, we found that increasing the gain of the support surface also altered normalized EEG-EMG coherence (main effect of Condition: F_2,1802.8_ = 6.027; p = 0.002). The coherence was greater for the medium difficulty condition when compared to low (medium vs. low mean difference=0.006; p = 0.011) and high (medium vs. high mean difference = 0.007; p = 0.007) difficulty conditions (Fig. 10). The coherence also modulated across frequency bands (main effect of Bands: F_4,1802_ = 4383.23; p < 0.001) and bins (main effect of Bins: F_6,1802_ = 13.60, p < 0.001). Importantly, across the stroke and healthy control group, there was modulation in normalized EEG-EMG coherence across difficulty conditions (significant Group × Condition interaction: F_2,1802.4_ =3.121, p = 0.044) and frequency bands (significant Group × Bands interaction: F_4,1802_ =15.013, p<0.001), but not across bins (no significant Group × Bins interaction: F_6,1802_ =1.678, p = 0.123; no main effect of Group: F_,1,18_ = 0.192, p =0.666; Fig. 10). None of the pairwise posthoc comparisons reached statistical significance (p>adjusted α for multiple comparisons).

**Fig. 10.**
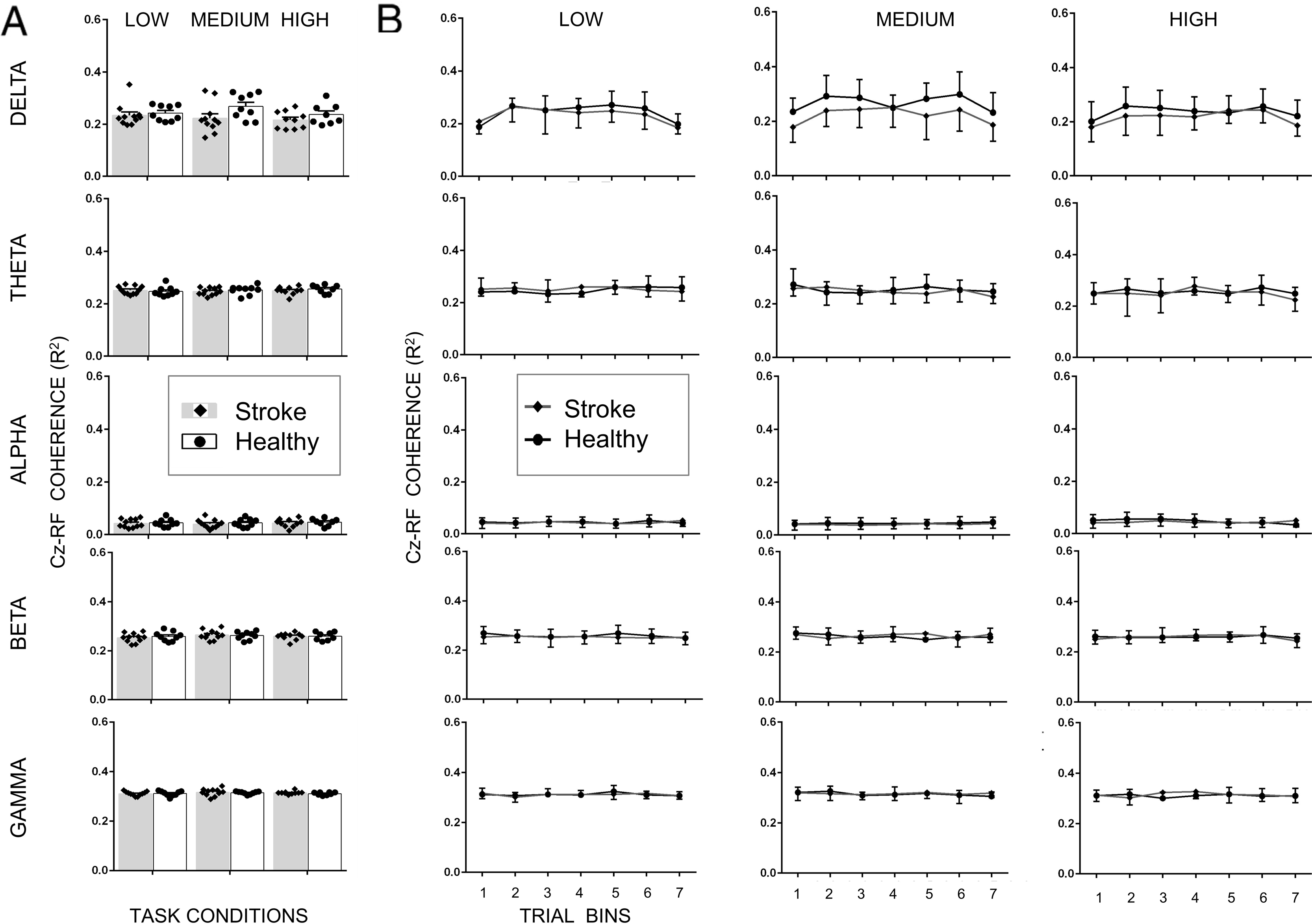
Cortico-muscular coherence (CMC) magnitude between Cz (EEG) and rectus femoris (EMG) for all five frequency bands and all three task conditions for stroke patients and healthy controls. **A.** CMC averaged across the entire 20 sec trial. **B.** CMC for seven trial bins. Data are means (± SEM) of all subjects. **P < 0.01, significant between-group differences.

## DISCUSSION

We found differences in corticomuscular coherence between stroke patients and healthy controls and this difference was dependent on the difficulty of the continuous balance task. Mainly, when compared with healthy control, stroke patients showed smaller magnitudes of coherence during the medium difficulty balance task condition for the distal leg muscles but not during quiet standing and high difficulty task condition. The group differences were primarily found at the delta frequency band but not at higher frequency bands.

### Lower corticomuscular coherence in the delta frequency band in stroke

Both stroke patients and healthy older adults showed significant delta band coherence between Cz and lower extremity muscles during the continuous balance task. An earlier aging study has reported presence of a significant coherence in the delta frequency band in both higher and lower leg muscles of healthy older adults during performance of a standing task under external perturbation (Ozdemir et al., 2018a). The delta band corticomuscular coherence during a sensorimotor task has been associated with the detection of salient stimuli for reactive control of movements (Novembre et al., 2018, 2019). Novembre and colleagues (Novembre et al., 2019) provided somatosensory or auditory stimulation to subjects performing an isometric force production task using the thumb and index finger. Following the stimulation, the authors observed the presence of delta band coherence between the motor cortex and force output. The occurrence of delta band coherence overlapped with negative and positive EEG potentials observed over the vertex region. This might suggest that the delta band corticomuscular coherence is associated with the cortical potentials in the region of vertex, although the cortical activation and corticospinal oscillations may have distinct mechanisms (M. R. Baker & Baker, 2003b). The delta band coherence was found to have a functional significance as it was associated with systematic force modulations during the isometric task (Novembre et al., 2019). The authors indicated that this coupling during the period of positive-negative EEG vertex potentials represents a detection of salient stimuli arising from unexpected events to modulate grip forces (Novembre et al., 2018, 2019).

During the balance task performed under challenging conditions, several studies have reported positive-negative EEG potentials in the frontocentral region (Dietz et al., 1984, 1985; Goel et al., 2018b, 2019c, 2021; Maki & McIlroy, 2007; Marlin et al., 2014; Mochizuki et al., 2010; Ozdemir et al., 2018a; Varghese et al., 2014, 2019). The negative EEG potential, which is also known as the N1 response, is argued to represent detection of an error (i.e., difference between the actual balance state due to balance challenge and the anticipated state) resulting in production of postural responses to the overcome the challenge. Our recent work using advanced computational techniques has identified presence of multiple EEG evoked negative potentials time-locked to moments of instability that may occur during a continuous balance trial (Goel et al., 2021). These EEG negative potentials have characteristics similar to those found during the platform perturbation task (Goel et al., 2018b; Ozdemir et al., 2018a). The delta band coherence might have occurred during moments causing the N1 response. However, the current analysis does not provide information on the relative timing of delta band coherence and EEG potentials. Future work will focus on understanding the significance of delta band coherence during challenging balance tasks.

We speculate that the presence of delta band coherence during the continuous balance task might represent detection of unexpected events in the form of instability caused due to somatosensory manipulation requiring a reactive involvement of lower leg muscles. Importantly, stroke patients showed lower magnitude of corticomuscular coherence in comparison to healthy adults when performing the task under the medium difficulty condition. The patients in our study experienced an MCA stroke which usually involves sensory and motor regions (Walcott et al., 2014). Thus, it is reasonable to expect impaired coordination and connectivity among sensorimotor networks involved in balance control (A. E. Edwards et al., 2018). Compromised context-dependent sensorimotor process might have led to significantly smaller delta band coherence in stroke patients. And the impact of this impairment is likely to be observed during the challenging balance task (mainly seen at the medium difficulty condition) where the gain of the support platform was altered resulting in continuous sensory manipulation of the balance control loop (Rasman et al., 2018). Subjects in both groups found the high difficulty condition equally challenging (i.e., similar number of falls), and thus we failed to observed a lack of group difference in coherence. The delta band coherence was smaller for all distal leg muscles but not proximal muscles on the affected side in stroke patients when compared with healthy controls. Similar changes in coherence for distal versus proximal muscles on the affected side was observed during elbow flexion and wrist extension tasks in chronic stroke patients (Mima et al., 2001). These findings might be due to different organization of pyramidal pathways to distal and proximal muscles (Grosse et al., 2002) or differences in recovery of the corticospinal connections or cortical reorganization (Mima et al., 2001).

### Similar coherence magnitude between stroke and healthy controls at higher frequency bands

We found significant coherence in the theta frequency band, but it was not different between stroke patients and healthy controls during the continuous balance task. The theta band coherence has been previously shown while standing and walking in healthy adults (Peterson & Ferris, 2019). These authors found that the theta band coherence increased with a pull perturbation during standing and suggested that coherence in the theta frequency band might be associated with increased muscle activity during perturbation. In our study, both stroke patients and healthy controls might have used similar levels of muscle activity during performance of the continuous balance task resulting in no difference in the theta coherence band.

Earlier work has shown that when subjects are instructed to keep their eyes closed versus open during a quiet standing task, the coherence in the alpha frequency band is significantly smaller (Vecchio et al., 2008). The alpha activity has been associated with the visuo-spatial attentional aspects of balance control (A. A. Edwards et al., 2021) and possibly reflect processing integrating visual and somatosensory information (Vecchio et al., 2008). In our study, all subjects were required to close their eyes during the continuous balance task that included two trials of quiet standing (low difficulty condition). We found that both stroke patients and healthy controls did not show coherence in the alpha frequency band (i.e., coherence values < Z). Our findings extend previous work by showing the absence of alpha band coherence during both quiet standing and standing on a sway-referenced platform with varying gain with eyes closed.

Although we found significant coherence magnitude in the beta frequency band during the continuous balance task, it was not different between stroke patients and healthy controls. The beta band coherence is usually observed during tasks involving sub-maximal or isometric muscle contraction (Gwin & Ferris, 2012; Ushiyama et al., 2012) and has been previously reported during standing tasks (Jacobs et al., 2015; Stokkermans et al., 2022). A recent study that instructed subjects to perform isolated ankle dorsiflexion found lower beta band coherence for lower leg agonist (TA) as well as antagonist (GM) muscles in stroke patients as compared to healthy control (Xu et al., 2023). The beta band coherence is believed to represent sensorimotor integration resulting from the interaction between motor cortex and peripheral structures such as spinal cord, muscles, and afferent nerves (S. N. Baker, 2007; Grosse et al., 2002). A lesion that affects the corticospinal connections as it happens during stroke would influence the beta band coherence (Mima et al., 1999, 2001). However, this was not the case in our study. Guo and colleagues reported similar findings where stroke patients showed unaltered beta band coherence when compared with healthy controls during performance of an upper limb isometric force contraction task (Guo et al., 2020). One possibility might be the functional status of stroke patients. In our study, stroke patients, although not fully recovered, had regained independence in performing daily activities such as standing and walking. Most stroke patients were able to stand on the balance platform for the entire duration of balance task without requiring a rest break or falling (except the high difficulty condition).

Similarly, we found no difference in the low gamma band coherence between stroke patients and healthy controls during the continuous balance task. The coherence in the gamma frequency bands has been observed during dynamic muscle contractions involving lower limbs (Gwin & Ferris, 2012) and while standing on a moveable platform (Stokkermans et al., 2022). The gamma band coherence is related to muscle dynamics, i.e., scales with the force produced by a muscle (Brown & Marsden, 1998; Mima et al., 1999). Although the continuous balance task that subjects performed involved both static (quiet standing or low difficulty) and dynamic movements (medium and high difficulty conditions), it failed to cause a modulation in the low gamma band coherence in both groups with the task difficulty. It is possible that the stroke patients in our study were in the chronic stage of recovery, having passed at least 12 months since their initial stroke, and had achieved a level of recovery such that they were able to produce dynamic muscle activities similar to healthy controls.

### Poor balance stability in stroke patients

Although stroke patients when compared with healthy controls showed poor scores on balance and mobility on clinical tests such as BB scale and TUG, we failed to observe a consistent difference across measures of balance performance as assessed on the computerized balance platform. Mainly, stroke patients showed higher RMS COP when compared to the healthy control, however similar difference were not observed for RMS COP velocity and COP path length. This finding is consistent with (Fujimoto et al., 2014b)who found improved BBS score after the balance training intervention in stroke patients but did not find consistent improvement in postural stability as assessed through COP sway and COP velocity. Higher RMS COP in stroke patients might be due to adoption of a postural control strategy that allows them to change movement direction and speed rapidly and purposefully at the cost of balance stability (Goel et al., 2019a). Such a strategy might help in adapting to the challenging conditions and has been observed in young adults (Goel et al., 2019a). However, in stroke patients this potential strategy led to the increased occurrence of falls (although nonsignificant) when compared with healthy controls during the most challenging balance task condition.

### Limitations

Considering the small sample size of this study, it is important to confirm these findings in a larger cohort. Our analysis for the corticomuscular coherence focused on the EEG sensor level activity which limits the spatial resolution to identify specific brain regions engaged during the balance task. It is generally believed that the EEG activity recorded using a sensor over the vertex region arises from brain areas in the frontocentral regions including anterior cingulate, supplementary motor area, primary motor cortex (Goel et al., 2018b, 2019a; Mouraux & Iannetti, 2009). Future work with advanced EEG source localization is needed to confirm the brain areas engaged during the continuous balance task. Lastly, it is recommended to recruit healthy participants who are age and gender matched to stroke patients.

### Summary and Conclusion

To our knowledge, this is the first study investigating cortico-muscular coherence in relation to balance control among stroke patients versus healthy controls with respect to all five frequency bands across distal and proximal leg muscles. Our finding of smaller delta frequency band corticomuscular coherence during the challenging continuous balance task in stroke patients when compared with healthy controls might suggest impairment in the context dependent sensorimotor processes for the detection of somatosensory modulation to control the postural muscles. Moreover, the observation of abnormal coherence for distal but not proximal leg muscles on the affected side further suggests differences in the (re)organization of the corticospinal connections across the two muscles groups for balance control.

## Data Availability

All data produced in the present study are available upon reasonable request to the authors

## ACKNOWLEDGMENT

This study was supported by a grant to PJP from the NIH National Center of Neuromodulation for Rehabilitation, the National Institutes of Health Eunice Kennedy Shriver National Institute of Child Health and Human Development (NIH/NICHD) under Grant P2CHD086844 and NIH/NICHD R25HD106896 to PJP. We would like to thank Drs. Charles Layne and Seoung Hoon Park for comments on an earlier version of the manuscript. We would like to thank Victoria Nacinovich, Sana Sheikh, Esther Jimenez, and Yoshua Carmona for helping with data collection. We would also like to thank Dr. Mauricio Ramirez, Michelle Patrick-Krueger and Alexander Steele for their help with technical issues and data analysis.

